# Understanding Unmet Needs and Opportunities for Digital Transformation in Nursing Admission Assessment: A Qualitative Study

**DOI:** 10.1101/2025.05.06.25327049

**Authors:** Yeyoon Choi, Junhyuk Seo, Hyun A Shin, Sook Hyun Park, Minjeong Park, Ye Eun Lee, Subin Jeon, Mijung Lee, Jeong Hee Hong

**Affiliations:** Department of Nursing, Samsung Medical Center, Seoul, Republic of Korea

**Keywords:** Nursing Admission Assessment, Nursing Workload, Documentation Burden, Digital Health, patient-centered Care, Qualitative Research

## Abstract

Nursing admission assessment is a critical first step in the inpatient care process, providing essential information for clinical decision-making and patient safety. However, despite its importance, this process places a significant burden on nurses, who face challenges such as time pressure, documentation overload, and communication barriers, particularly among elderly patients. This qualitative descriptive study explores the experiences of nurses, patients, and caregivers regarding the nursing admission assessment process at a large tertiary hospital. Through focus group and in-depth interviews, we identified four major themes: (1) perceived meaning of the assessment, (2) operational and systemic challenges, (3) communication challenges among stakeholders, and (4) suggestions for future improvement. Based on these findings, we propose the development of a self-report questionnaire, a digital pre-admission system, enhanced EHR integration, and user-centered system design. These insights offer guidance for workflow innovations aimed at reducing nursing workload while maintaining high-quality care.

## 1. Introduction

Nursing admission assessment is an essential component of inpatient care, serving as the cornerstone of the nursing process [1]. Through activities such as reviewing medical records and conducting physical evaluations, nurses establish the foundation for all subsequent nursing interventions, evaluations, and care planning decisions [2]. Conducted at the outset of the patient journey, the admission assessment provides a comprehensive clinical overview[1], enabling the identification of care needs and potential risks, thereby enhancing patient safety [3]. Crucially, the value of this process lies in the nurse’s clinical expertise, which allows for the tailored understanding of each patient’s unique condition and appropriate assessment. Building on the foundational role of admission assessments, subsequent nursing assessments throughout the care process have been shown to be indispensable for detecting deterioration and improving patient outcomes [4, 5, 6]. Furthermore, the concerns and clinical impressions developed in the process are now recognised as strong predictors of patient outcomes beyond physiological data [7, 8].

Despite its vital role, nursing admission assessment imposes a considerable burden on nurses. The assessment requires high levels of data literacy and critical thinking, involving the reconstruction, exploration, documentation, and communication of patient data [2]. It is also crucial to complete the assessment in a timely manner, as it gives up-to-date references for healthcare delivery and decision-making [9, 10]. A core component of admission assessment is the use of electronic health record (EHR) and documentation, which often demands extra time and effort [11]. Over time, the burden of EHR-related documentation has emerged as a major contributor to nurse burnout [12, 13]. Following the global adoption of EHRs has transformed them into essential systems for healthcare delivery, encompassing functions such as prescriptions, workflows, and communication [14, 15]. This integration has enhanced patient safety [16] and has also provided extended opportunities for patients and providers—including personalized care, data-driven research, and clinical decision support systems [17, 18, 19, 20, 21]. Nevertheless, many papers emphasize EHR-associated burnout among nurses and physicians, which may lead to negative patient outcomes [22, 23, 24, 25].

While recognition of documentation burden is increasing, efforts to improve the efficiency of nursing admission assessments remain limited. One prior study demonstrated that optimizing nursing admission documentation reduced documentation time by 34% [12]. Similarly, multiple studies have consistently identified EHR usability as both a contributing factor to burnout and a potential area for improvement [24, 26]. However, this technological problem is not the only factor burdening nurses. A recent research suggests that relieving documentation burden should consider non-EHR factors—staffing, patient acuity and complexity, and efficient time management [27]. These multifaceted challenges require comprehensive solutions that go beyond simple interface improvements. Recognizing that simple adjustments to EHR interfaces cannot fully resolve the documentation burden, researchers and practitioners have explored broader digital health strategies to redesign clinical workflows. These innovations such as voice-based documentation, barcode systems, and artificial intelligence (AI) have been proposed to relieve the burden on providers [28, 29, 30, 31]. Furthermore, both the World Health Organization (WHO) and International Council of Nurses (ICN) emphasized digital health’s role in addressing these challenges [32, 33]. Therefore, redesigning the future of nursing admission assessment is now imperative.

In response to this need, we conducted a qualitative study to explore opportunities for improving the efficiency of nursing admission assessment. This study had two primary aims: to explore the current admission journey to understand the expectations and limitations of key stakeholders—nurses, patients, and caregivers; and second, to investigate ways to create a more efficient admission assessment process. Through a qualitative descriptive study, we identified key pain points and opportunities to improve the workflow. Uniquely, our study integrates perspectives from end-users, capturing the full scope of the admission assessment interaction.

## 2. Methods

### 2.1. Study design

A qualitative descriptive study design was employed to explore how stakeholders experience the nursing admission assessment process. This approach is widely used for examining clinical phenomena [34], as it captures direct and detailed accounts of participants’ experiences [35]. Moreover, qualitative descriptive studies are particularly useful for laying the groundwork for future intervention development [36]. Using “who,” “what,” “where,” and “why” questions, we conducted an in-depth investigation of the nursing admission assessment.

Focus group interviews (FGIs) with nurses and in-depth interviews with patients and caregivers were conducted. This differentiated approach reflected the distinct nature of their experiences. FGIs allowed nurses to share challenges collaboratively, whereas individual interviews ensured patient privacy.

### 2.2. Recruitment

Participants were recruited between January and March 2025 at a large tertiary hospital in Seoul, South Korea. During recruitment and interviews, the primary researchers (YC, JS) screened all interested individuals, assessed eligibility based on predefined criteria (Table 1), and obtained written informed consent.

**Table 1:**
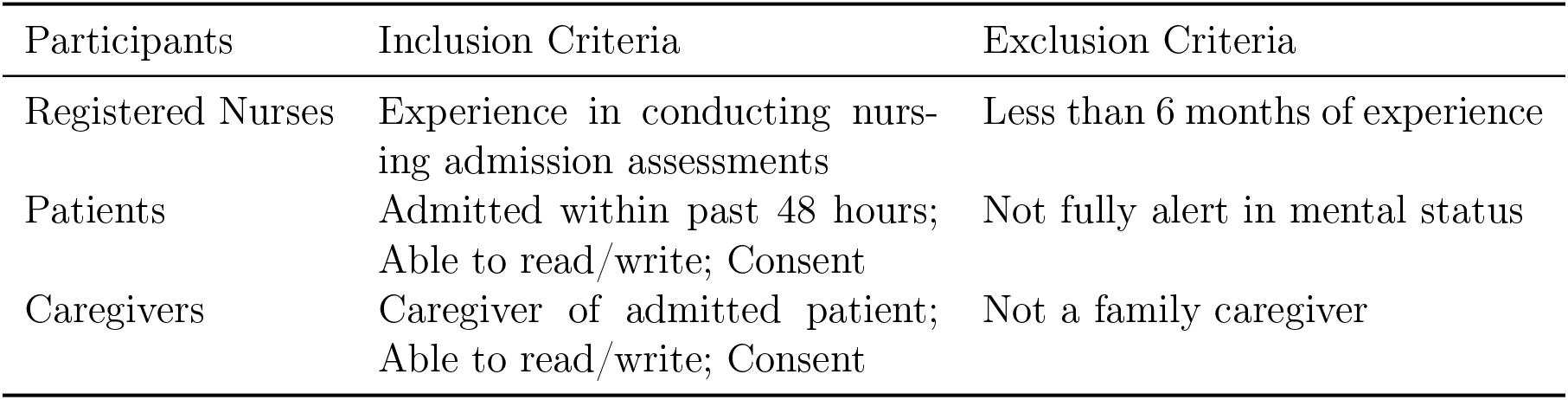
Eligibility Criteria of Participants.

| Participants | Inclusion Criteria | Exclusion Criteria |
| --- | --- | --- |
| Registered Nurses | Experience in conducting nursing admission assessments | Less than 6 months of experience |
| Patients | Admitted within past 48 hours;<br>Able to read/write; Consent | Not fully alert in mental status |
| Caregivers | Caregiver of admitted patient;<br>Able to read/write; Consent | Not a family caregiver |

Registered nurses were recruited through purposive sampling to ensure diverse representation across experience levels and clinical departments. Recruitment was announced via posters placed on hospital notice boards outlining the eligibility criteria. A total of eight nurses (n = 8) were recruited and divided into two focus groups based on clinical experience: senior nurses (>=4 years of experience, n = 4) and junior nurses (<4 years of experience, n = 4). Nurses were selected from a variety of clinical units, including pulmonary, oncology, cardiology, and rehabilitation wards, to ensure a broad perspective on the nursing admission assessment process.

Patients and caregivers were recruited from pulmonary and oncology wards. Six individual patient interviews (n=6) and six joint interviews with patients and caregivers (n=6 each) were planned. Based on early findings, caregiver participation was added after IRB approval.

### 2.3. Data collection

Data were collected through semi-structured interviews guided by pre-developed interview guides. The guides were developed by six researchers to ensure clinical relevance and depth. The research team was composed of registered nurses from various units (including Pulmonary, Oncology, Neurosurgery, and Psychiatry), all of whom were clinical nurses with more than four years of experience.

The FGIs with nurses focused on three main areas: (1) current practices and challenges in nursing admission assessment, (2) challenges in patient interactions, and (3) recommendations for improving the admission process. Each FGI lasted approximately one hour and was conducted by two interviewers—a primary (HS) and a secondary (YC). The primary interviewer had prior qualitative research experience at the doctoral level, and both had completed formal coursework in qualitative methods. An additional researcher (JS), who had also undergone the coursework, attended each FGI as an observer, taking field notes and monitoring group dynamics. The observer documented non-verbal cues and interpersonal interactions that might not be captured in audio recordings.

After interviewing nurses, we defined three domains to explore with patients: (1) Recent experience with nursing admission assessment, and (2) Recommendations for improving the admission process and required features. Individual in-depth interviews with patients lasted approximately 20 to 30 minutes and were conducted in a private setting. The observer was also present during these interviews, and verbal consent for observation was obtained from each participant. All interviews were audio-recorded and transcribed verbatim within 48 hours by the interviewer. Transcripts were then reviewed and annotated by the observer to capture nuances of interaction and context.

### 2.4. Data analysis

Transcripts were analyzed using MAXQDA (v24.8.0) tool [37], following Braun and Clarke’s six-phase thematic analysis[38]. A hybrid approach combined deductive coding (aligned with study objectives) and inductive coding (emergent themes). Two researchers independently coded transcripts and resolved discrepancies through discussion, with broader team consensus.

Themes were validated using triangulation across nurses, patients, and caregivers.

### 2.5. Rigor

To ensure rigor in our study, we applied Lincoln and Guba’s (1985) four criteria[39]. Additionally, we incorporated Muthanna and Alduais’s (2023) reflexivity framework to maintain vigilance regarding researchers’ positions and actions throughout all stages of the research process[40]. (Table. 2)

**Table 2:** Methodological and Analytical Rigor.

| Quality Criteria | Action taken by the researchers |
| --- | --- |
| Credibility | - Research team comprised of registered nurses with over 4 years of clinical experience<br>- Iterative discussions among co-authors to review codes and themes<br>- Data source triangulation across nurses, patients, and caregivers<br>- Independent coding by two researchers |
| Dependability | - Purposive sampling across diverse clinical units<br>- Individual investigation of participant subgroups<br>- Detailed description of participant characteristics and clinical setting |
| Confirmability | - Transparent description of methodology and coding process<br>- Use of MAXQDA and Braun & Clarke's six-phase analysis |
| Transferability | - Use of direct participant quotes<br>- Audit trail and documentation of analytic decisions<br>- Reflexive discussion among researchers |
| Reflexivity | - Reflexive journaling during research<br>- Disclosure of researchers' clinical backgrounds<br>- Iterative discussions with research team to mitigate potential bias |

### 2.6. Ethics

The study was approved by the Institutional Review Board of Samsung Medical Center (IRB No. 2024-11-019). Participants provided written informed consent and received a small token of appreciation.

## 3. Results

### 3.1. Participants characteristics

Participants included seven nurses, five patients who were individually interviewed, and three patient-caregiver pairs. In total, 18 individuals participated (n = 18; nurses = 7, patients = 8, caregivers = 3).

Among the nurses, although eight were initially recruited, one senior nurse withdrew due to scheduling conflicts, resulting in seven participants. As shown in Table 3, there were clear distinctions between the junior and senior groups: all junior nurses were in their twenties with less than four years of experience, whereas the senior group showed greater diversity in both age and career length.

**Table 3:** Participant Characteristics of Registered Nurses.

| Characteristics | Junior Group (n=4) | Senior Group (n=3) |
| --- | --- | --- |
| **Age, n (%)** |  |  |
| 20–29 | 4 (100.0) | 1 (33.3) |
| 30–39 | 0 (0.0) | 1 (33.3) |
| 40–49 | 0 (0.0) | 1 (33.3) |
| **Sex, n (%)** |  |  |
| Female | 4 (100.0) | 3 (100.0) |
| **Clinical Experience, n (%)** |  |  |
| 1–3 years | 2 (50.0) | 0 (0.0) |
| 3–5 years | 2 (50.0) | 1 (33.3) |
| 5–7 years | 0 (0.0) | 1 (33.3) |
| ≥10 years | 0 (0.0) | 1 (33.3) |

For the patient and caregiver groups, five patients were interviewed individually, and three patient-caregiver pairs were recruited. Recruitment was discontinued after data saturation was reached, as no new concepts emerged during the final interview. The characteristics of patients and caregivers are presented in Table 4. More than two-thirds of the participants were elderly patients aged over 70, reflecting the general demographics of inpatient admissions. The variety of patients’ educational levels and numbers of previous hospitalisations enriched the context of the research.

**Table 4:**
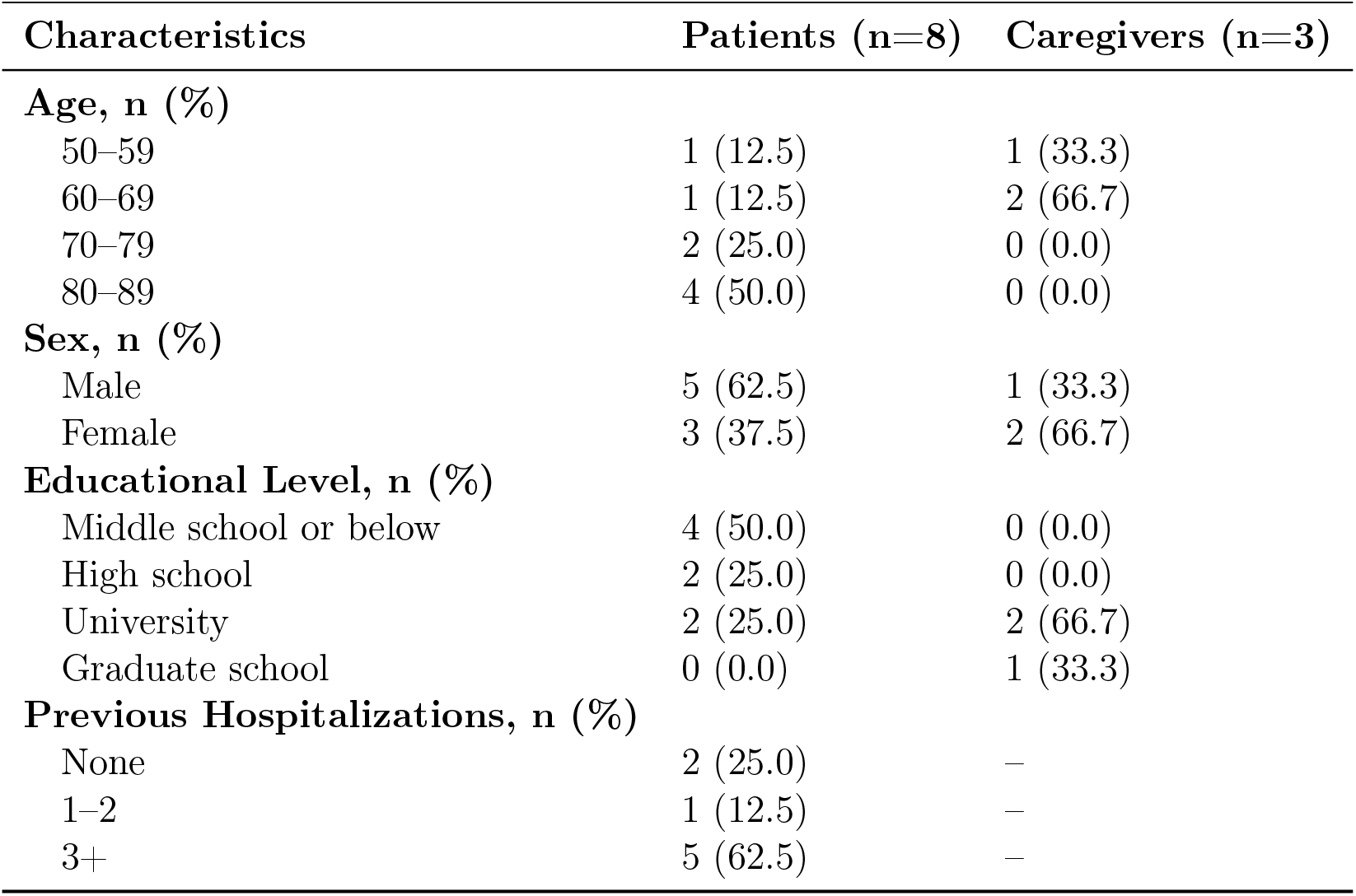
Participant Characteristics of Patients and Caregivers.

| Characteristics | Patients (n=8) | Caregivers (n=3) |
| --- | --- | --- |
| **Age, n (%)** |  |  |
| 50–59 | 1 (12.5) | 1 (33.3) |
| 60–69 | 1 (12.5) | 2 (66.7) |
| 70–79 | 2 (25.0) | 0 (0.0) |
| 80–89 | 4 (50.0) | 0 (0.0) |
| **Sex, n (%)** |  |  |
| Male | 5 (62.5) | 1 (33.3) |
| Female | 3 (37.5) | 2 (66.7) |
| **Educational Level, n (%)** |  |  |
| Middle school or below | 4 (50.0) | 0 (0.0) |
| High school | 2 (25.0) | 0 (0.0) |
| University | 2 (25.0) | 2 (66.7) |
| Graduate school | 0 (0.0) | 1 (33.3) |
| **Previous Hospitalizations, n (%)** |  |  |
| None | 2 (25.0) | – |
| 1–2 | 1 (12.5) | – |
| 3+ | 5 (62.5) | – |

### 3.2 Thematic overview of qualitative data

Four overarching themes emerged from the data (Table 5), illustrating perceived realities, reflecting current challenges, and presenting possible solutions in the nursing admission assessment process. Supplementary File 4 provides an overview of the coding framework.

**Table 5:** Themes and Sub-themes Identified in the Study.

| Theme | Sub-themes |
| --- | --- |
| Perceived meaning of Nursing Admission Assessment | - Data-driven and comprehensive assessment<br>- Starting point of holistic care |
| Operational and systemic challenges in Nursing Admission Assessment | - Managing time pressure with a self-report questionnaire<br>- Emotional burden under unpredictable admissions<br>- Documentation burden and systemic inefficiencies |
| Communication challenges among stakeholders | - Challenges in communicating with elderly patients<br>- Essential role of caregivers<br>- Misunderstandings of medical terminology and questionnaire |
| Suggestions for future improvement | - Digital pre-admission system<br>- System design accommodating stakeholders' needs<br>- Concerns about digital transformation of Nursing Admission Assessment |

### 3.3. Perceived Meaning of Nursing Admission Assessment

#### 3.3.1. Data-driven and comprehensive assessment

The nurse participants considered the nursing admission assessment to be not just a questioning or examination, but rather a comprehensive process that begins before the patient’s admission. Especially in their dedicated duty of admission management, tasks such as reviewing outpatient documents, historical data of admissions, and drug use are regarded as core assignment.

> “I thoroughly review all outpatient charts before meeting a patient. I think that time is really about gathering all scattered data. Sometimes I spend up to one or two hours before meeting patients just reviewing outpatient charts.” (Junior nurse 4)

While progress notes were documented by various medical departments from their individual perspectives, nursing assessments serve to unify these viewpoints. By integrating different medical perspectives into a single comprehensive view, these assessments create a cohesive foundation for understanding patients holistically and establishing the clinical context.

> “When looking at the EHR, there are so many outpatient records that I have to click through and check each one. We have to figure out which departments they visited, whether they’re still going there, and we even have to organize all their diagnoses ourselves.” (Senior nurse 1) Overall, these assessment procedures go beyond mere questioning and recording—they serve as a concrete process of clinical evaluation. Through systematic data collection and interpretation, the initial assessment creates the essential foundation for comprehensive clinical understanding.

#### 3.3.2. Starting point of holistic care

Senior nurses, more so than junior nurses, repeatedly emphasized that the nursing admission assessment should serve as a cornerstone of a holistic nursing plan, which covers head-to-toe, front-to-back patient care. Even though it is difficult to perform the procedure perfectly, the nurses understood that the original intention of the assessment is meant to be a continuous and comprehensive flow.

> “I feel burdened by the responsibility of having to include everything from the medications they’re taking to a full head-to-toe assessment, and even determining what prescriptions might be needed.” (Senior nurse 1)
>
> “As we learned in nursing school, there’s this inherent concept that during admission, we need to assess all body systems comprehensively… It would be ideal to consider the patient’s living environment after discharge and their activity levels, but… in reality, we’re so busy that we can’t pay as much attention to those aspects as we should.” (Senior nurse 2)

This perception of the nurses regarding admission assessment aligned with the attitudes of patients and caregivers. Some patient participants acknowledged that the nurses’ detailed understanding of their information was required to provide better care, and they made an effort to share information straightforwardly. Moreover, they had little concern when providing their sensitive data, even when others were nearby: “Since everyone goes through the same experience when hospitalized… I should just share everything without holding anything back.” (Patient 4)

Like the patients, caregiver participants also recognised the need for the assessment and understood that it should collect accurate information about the patients they care for. This suggests that patients and caregivers viewed the admission process as a natural part of the care journey rather than a burdensome task. One caregiver defended the thorough admission assessment as follows “It is because they should properly understand the patient’s information.” (Caregiver 8)

As remarked above, nursing admission assessment is regarded as an essential and comprehensive procedure for all stakeholders in the patient admission journey.

### 3.4. Operational and systemic challenges in Nursing Admission Assessment

#### 3.4.1. Managing time pressure with a self-report questionnaire

The initial interview question for nurses, patients, and caregivers asked them to recall their experiences with nursing admission assessments. A distinctive feature of the process was the self-report questionnaire on paper. Many general wards in the study hospital had developed their own forms tailored to their typical patient populations. The nurses considered this paper form essential for managing their time pressures. They described how a structured workflow was followed to streamline the process (Figure 1).

**Figure 1.**
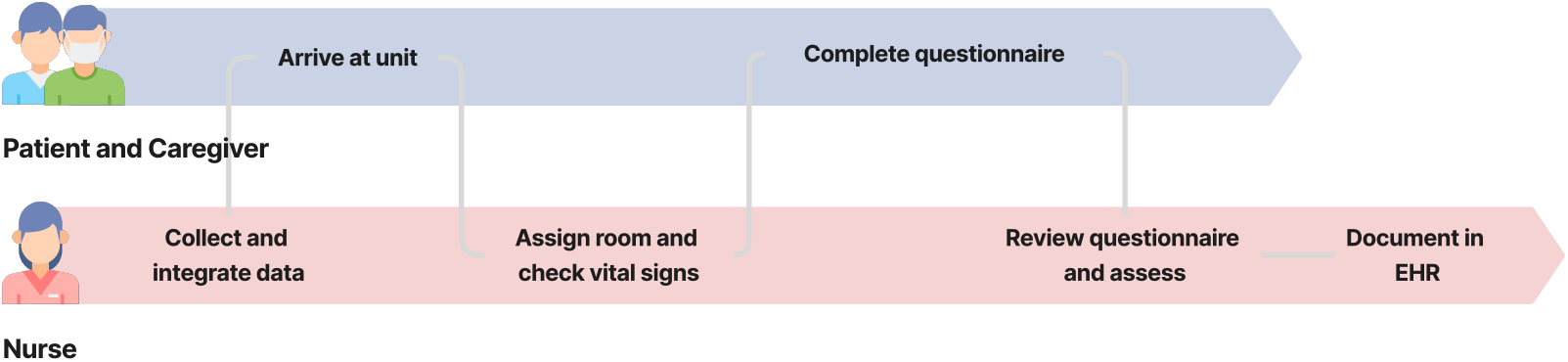
Process of typical nursing admission assessment at the study hospital.

Although not formally outlined in the protocol, this process was widely used across units for patients admitted for scheduled procedures or predictable treatment courses. Nurses routinely employed it to streamline workflow, but it also introduced potential complications. When the self-report questionnaire was incomplete, nurses had to revisit patients multiple times, causing disruptions and delays: “Sometimes, I collect the questionnaire first and enter them later, only to realize that there are gaps. Then, I have to go back and ask the patient again… “ (Junior nurse 2)

To manage time pressures, some nurses pre-filled patient data using EHR records before actually meeting the patient, confirming details with closed questions rather than conducting in-depth assessments. While this approach helped balance efficiency and feasibility, nurses remained concerned about data reliability and the depth of assessment.

> “I try to get as much information as possible from the chart beforehand. Then, when I meet the patient, I just confirm what I already know. The conversation ends up being more about fact-checking rather than a real interaction.” (Junior nurse 1)
>
> This perception of time pressures manifested in the nurses’ behavior, at times making patients feel uncomfortable. While patients wanted the nurses to slow down and explain things more carefully, the nurses felt too pressed for time to do so.
>
> “We don’t have enough time for thorough patient interviews, so we end up cutting conversations short. Even if a patient wants to share their worries, we have to move on quickly.” (Junior nurse 1)
>
> “Their explanations seem like they’re just mechanically rattling things off due to routine - it’s not just talking, but more like they’re just spitting out words… That’s how it feels, and for me, when there’s unfamiliar information, I wish they would slow down a bit… But it’s not that this person is doing it on purpose, it’s because they’ve developed this habit of speaking so rapidly.” (Patient 3)

While nurses are encountering this time pressures, caregivers of elderly patients also face time constraints during the admission process. Both patients and caregivers experience pressure from the tight time frames, disappointing their satisfaction with the journey.

> “Since we don’t live together anymore, we need to ask questions. So when we have to do this suddenly, we need to ask father in a rush, but if we could do it in advance, we could ask and prepare the medications ahead of time and write everything down like this. Often father doesn’t even know what medications he’s currently taking… “ (Caregiver 6)

#### 3.4.2. Emotional burden under unpredictable admissions

Nurses consistently reported that nursing admission assessments created a significant burden. This was especially evident when nurses take P-shifts (10am-7pm), which dedicated to manage most of the patient admissions in the study hospital. Given the high volume of patient admissions, assessments had to be conducted rapidly, often within 30 to 90 minutes per patient, depending on complexity. When multiple admissions occurred simultaneously, nurses found themselves rushing through assessments, leading to limited patient engagement and increased stress. Many nurses reported that P-shifts, involving patient admission management, remained burdensome despite their increased experience. Both junior and senior nurses emphasized that the unpredictability of the shift and the pressure to complete assessments quickly remained a major source of stress, regardless of experience level: “Even after five years, P-shifts are still stressful. Every day is different, and I never know how many patients I’ll need to assess.” (Senior nurse 1)

Due to unpredictability and high workload, nurses often rushed through assessments, leading to feelings of guilt and frustration about their inability to provide more comprehensive patient care: “I try my best, but sometimes I feel guilty. I wish I could spend more time with each patient, but there’s just no time.” (Junior nurse 3)

Despite the nurses’ frustrations, most patients expressed satisfaction with the nurses and their assessment processes. “…they are all very kind and thorough, without leaving anyone out.” (Patient 1), “They frequently come to check on me and explain everything about my schedule, so I have no complaints. I’m just doing what I’m told to do.” (Patient 4) This suggests that nurses are successfully maintaining quality of care despite their internal struggles during the patient journey.

#### 3.4.3. Documentation burden and systemic inefficiencies

Even when using the self-report questionnaire, entering admission assessments into the EHR presents a significant additional burden for nurses. According to one nurse participant, documentation tasks consumed over half the time spent on admission assessments, with this inefficient process taking away from direct nursing care:

> “I felt like about half my time was being lost. I wanted to provide more direct nursing care, but I had to fill out the EHR… The process of collecting information was so inefficient that it was very frustrating.” (Junior nurse 1)
>
> When there are multiple admissions, the repetitive task of having to input initial assessments for each patient was often perceived as a significant workload burden. This process became even more time-consuming depending on the structure of assessment items and input methods, highlighting the need for more efficient alternatives.
>
> “…I actually prefer transfers between units because we don’t have to do initial assessments for them. But for new admissions, even if they’re simple cases, we have to input everything one by one which takes a long time…” (Senior nurse 1)

Despite increasing digitalization of healthcare in South Korea, a critical nation-wide challenge persists: the inability to integrate patient records across different hospitals. This systemic limitation extends beyond individual hospital capabilities, reflecting a broader national healthcare issue. Consequently, nurses across the country must engage in time-consuming data compilation and direct hospital-to-hospital communication to gather comprehensive patient information.

> “Although outpatient charts contain admission history and medication records from other hospitals, we have to manually re-enter everything in the ward. I wish there was technical integration.” (Junior nurse 4)

This lack of data integration was particularly burdensome in managing patients’ personal medications. Nurses spent considerable time organizing and verifying medications when there were significant differences between medications brought by patients and those in prescription. Furthermore, patients often failed to bring their medications even they were told to. This lack of data integration resulted in inefficiencies in nursing admission assessments and increased the workload burden on nurses.

> “…I once spent two hours just organizing personal medications. The medications were so diverse, different from prescriptions, and mixed together, which took a lot of time to sort out.” (Senior nurse 2)

### 3.5. Communication challenges among stakeholders

#### 3.5.1. Challenges in communicating with elderly patients

Nurses reported that collecting health information from elderly patients is one of the most challenging aspects of the admission assessment. Many old patients struggle to clearly explain their medical history and current medications. They also hardly recall their past operation history, diagnosis dates, reasons for taking specific medications, and other important medical details.

> “When we ask ‘Are you taking any medications?’ they show us this much medicine, but they don’t know why they’re taking them. They just say they take them because the hospital told them to.” (Junior nurse 4)

Impaired hearing and vision in elderly patients pose additional challenges for nurses during assessment. Nurses must carefully document responses while asking numerous follow-up questions to gather complete information. This communication barrier leads to prolonged assessments and increases the nurses’ workload: “There were cases where patients had poor hearing and couldn’t write. For those patients, it took over 30 minutes just to complete one question.” (Junior nurse 3)

#### 3.5.2. Essential role of caregivers

Nurses reported that caregivers serve as crucial information providers during nursing admission assessments. Particularly for elderly patients, caregivers’ knowledge of patients’ medical history and medications helped provide more accurate information about patients’ health status. However, some elderly caregivers did not have complete knowledge of patients’ health information.

> “When caregivers are young and present to assist patients, the assessment is often nearly perfect…However, when the caregiver is an elderly spouse, it can be difficult to obtain accurate information.” (Senior nurse 2)

Nevertheless, the presence of caregivers played a crucial role in the nursing admission assessment process. When caregivers were present, the accuracy of information improved, and the assessment process became shorter: “It’s much better when their caregivers are present. It takes much less time.” (Senior nurse 3)

In conclusion, caregivers acted as essential information providers for nurses, significantly improving the quality and efficiency of admission assessment. However, the accuracy of detailed information could vary depending on the caregiver’s age and health literacy, requiring nurses to conduct additional verification processes to supplement the information in such cases.

#### 3.5.3. Misunderstandings of medical terminology and questionnaire items

Nurses reported that patients often struggled to understand the admission assessment items. In particular, since the medical terminology used in the assessment questions did not align with patients’ everyday language, nurses needed to “translate” these terms into more understandable expressions.

> “Since what we’re trying to ask here differs from what patients understand, I need time to ‘translate’ it for them.” (Junior nurse 4)

To bridge this gap, nurses had to translate assessment items into patient-centered expressions. For example, instead of asking “Do you have chest pain?”, they would rephrase it as “Do you feel any tightness or squeezing sensation in your chest?” While this approach made questions more understandable, it significantly increased the time spent on assessments and added to nurses’ workload. Additionally, there were concerns about whether the intended meaning of assessment items was being accurately conveyed.

> “For example, when a patient checks ‘sleep disorder’ and I ask ‘Have you always had trouble sleeping?’, they might say ‘I only slept 6 hours this past week because I was anxious about admission.’ However, this is not something I can readily classify as a sleep disorder.” (Junior nurse 4)

As a result, nurses had to explain the intended meaning and adapt responses to reconcile discrepancies between patients’ understanding and assessment items, leading to longer assessment times. This additional work became especially burdensome when nurses needed to assess multiple new patients.

A significant issue with the self-report questionnaire on paper was that patients often skipped unclear items rather than asking for clarification. While most patients consulted their caregivers about confusing items, one participant admitted to marking answers randomly: “There were some things I didn’t know. Maybe once or twice. I just checked them randomly.” (Patient 5)

This comprehension challenge extended beyond paperwork to face-to-face interviews, suggesting that the questionnaire items themselves needed to be more patient-friendly regardless of the assessment method: “I don’t have any shortness of breath, cough, or phlegm, so that’s fine, but what does arrhythmia mean?” (Patient 1)

### 3.6. Suggestions for future improvement

#### 3.6.1. Digital Pre-admission system

During interviews with junior nurse participants, we encountered a significant insight that suggested a potential way to reduce nurses’ workload. The nurse firmly stated that a pre-admission self-report system would make this process much easier.

> “One patient was a nurse who had prepared and brought in her medical history and health information in advance. It was incredibly helpful… If patients enter their information in advance, it would definitely save more than 10 minutes. I’m confident about that.” (Junior nurse 3)

Based on this case, when the senior nurses were asked about the potential impact of implementing a system where patients could enter their health information in advance, most responded positively. Many nurses suggested that if patients could input key health information through pre-admission screening and link it to the EHR system, it would reduce uncertainty and accelerate the assessment process from the nursing perspective. One senior nurse elaborated: “If patients could input their information in advance and we just verify any missing parts, it would make our work so much easier.” (Senior nurse 3)

Patients also indicated that being able to prepare their information ahead of time would be far more beneficial than having to provide it at the time of admission: “We really appreciate being given this kind of preparation in advance. If we get it 4-5 days before, I think all patients would find it helpful.” (Patient 1)

Additionally, some patients and caregivers acknowledged the practicality of a mobile questionnaire, recognizing that digital transformation is becoming a natural progression in the hospital admission process: “Shouldn’t we move towards mobile solutions in the future? … Using mobile would save time and such. I think this level would be manageable.” (Patient 8)

#### 3.6.2. System design accommodating stakeholders need

Nurses emphasized that the success of the digital pre-admission system would depend on questionnaire design—items needed to be easy for patients to complete while clearly indicating which elements required direct nursing assessment. This balanced approach would enhance patient participation and system practicality.

> “Basic information like occupation, marital status, primary caregiver, and allergies can be filled out by patients in advance. On the other hand, things like pressure ulcers or fall risks need to be directly assessed by us, so it would be good to structure the system to distinguish between these types of information.” (Senior nurse 2)

Nurses expected that linking patient-entered information to EHR would reduce assessment time, but emphasized the need for nurses to be able to verify and modify the information: “It would be good if patients’ responses could be linked to the system. And I think we need a process where we can review it once.” (Senior nurse 2)

As a result, nurses expected that the digital pre-admission system would improve the efficiency of the initial nursing assessment and encourage active information provision from patients. However, they emphasized that it would be essential to design questionnaire items to be easily understood by patients and to include a nursing verification process to review the reliability of entered data.

Based on our in-depth interviews with patients and caregivers, we identified five essential requirements for implementing an effective digital pre-admission system: **ensuring visual accessibility** through sufficiently large fonts and clear readability features to accommodate elderly patients; **simplifying medical terminology** into everyday language that patients can easily understand, such as using “blood pressure medicine” instead of specific pharmaceutical names; **designing concise and focused questionnaire** to maintain patient engagement and prevent incomplete responses; **providing clear advance guidance** to help users understand both the purpose and significance of the pre-admission process; and **offering flexible input methods** that allow users to choose their preferred way of entering information based on their comfort level and technological proficiency.

Participant feedback strongly supported these requirements:

> “It would be better if the text could be bolder and clearer.” (Patient 4)
>
> “We can’t use specific medication names—we can only write things like ‘blood pressure medicine’ or ‘diabetes medicine’.” (Caregiver 6)
>
> “If it’s too long, people will just give up.” (Patient 7)
>
> “If they tell us in advance what content will be on mobile, we’ll answer sincerely. But if it comes without any explanation, we might not do it.” (Patient 8)
>
> “Voice input is fine, but if they tell us to touch, we can do that too.” (Patient 4)

Thus, patients and caregivers emphasized the importance of being able to understand and respond accurately to questions, rather than just the technical possibility of ‘being able to fill it out digitally’.

#### 3.6.3. Concerns about digital transformation of Nursing Admission Assessment

While nurses generally expressed positive attitudes towards the digital pre-admission system’s potential to improve the efficiency of initial nursing assessments, one nurse raised concerns about the possibility of increased burden on nurses due to excessive patient responses.

> “I’m worried that some patients might write too much if we provide a preadmission questionnaire. For example, if we don’t specify to only write symptoms ‘within the past month,’ some people might write something like a research paper. Then it could have the opposite effect of our intention to reduce nurses’ workload.” (Junior nurses 3)

Additionally, one nurse mentioned the emotional burden of introducing a new system into clinical practice. Even if the computerization of nursing admission assessment and digital pre-assessment system are designed as efficient tools, there was a concern that nurses in the field might find it unfamiliar and be hesitant to accept it: “It’s not easy to accept things like this. Sometimes people think ‘here they go again with something strange…” (Senior nurse 1)

One the other hand, elderly patients and their caregivers, in particular, not only showed a lack of confidence in operating mobile devices but also wariness of external threats such as voice phishing scams: “These days, I just hang up if it’s an unknown number. There are so many voice phishing scams lately…” (Patient 1)

Additionally, some patients expressed concerns about making mistakes during the information entry process, given that the accuracy of questionnaire responses significantly impacts their treatment. They also mentioned that when there are too many questions or unfamiliar medical terms, patients might skip responses or fill them out arbitrarily due to lack of understanding: “If there’s a word I don’t know, I’ll just skip it. I’m not going to bother trying to check those things…” (Caregiver 6), “Since we’re coming here with a serious illness, I’m worried about making even a small typo or mistake.” (Patient 7), “It’s better to ask directly. That’s more comfortable and accurate.” (Caregiver 8)

Beyond technical ability, it became apparent that patients did not perceive completing digital questionnaires as part of their responsibilities during admission. “…that’s not going to be easy. Especially since accessing things on mobile isn’t really seen as something we’re obligated to do.” (Patient 8). One caregiver specifically emphasized the need for a reward system.

> “To activate something like this, there needs to be some benefit… Rather than financial rewards, for example with admission - if you do this on mobile, you get priority in room assignments. If you get priority, that would be good. If you give rewards like this, people will definitely use the mobile system.” (Caregiver 6)

These statements suggest a potential gap between users’ digital capabilities and system design in nursing admission assessments. One patient participant emphasized the value of direct communication, noting that while digital-based assessments may have fewer errors, they lack the human warmth present in face-to-face interactions: “Since it’s human-to-human interaction, there are mistakes. Because humans are human. But while AI doesn’t make mistakes, it’s cold and rigid. That’s the difference.” (Patient 3)

## 4. Discussion

### 4.1. Implications

#### 4.1.1. Identified importance and barriers in Nursing Admission Assessment

Our findings revealed that both nurses and patients recognised the importance of nursing admission assessments. Nurses viewed them as critical for care planning and safety monitoring, while patients demonstrated strong engagement, seeing the assessment as an opportunity to share their health concerns and reflect on their overall health status. Patients expressed gratitude for the thorough questioning and demonstrated active participation in providing detailed information.

However, nursing admission assessment is described as a task that places a heavy workload on both junior and senior nurses. Both groups emphasised how the uncertainty surrounding patients’ conditions creates a significant time burden, yet they must complete these assessments quickly during their shifts. This was identified as a key reason why nurses cannot conduct thorough assessments and patient interviews. Most nurses expressed concerns about incomplete admission assessments, which might compromise patient safety.

Interestingly, although many nurses expressed concern about providing incomplete assessments due to time pressure, most patients reported high levels of satisfaction with their care. This discrepancy between nurses’ internal perceptions and patients’ experiences is a key insight, highlighting the importance of developing solutions that support nurses in upholding their professional standards without imposing excessive self-blame or stress. Rather than lowering care expectations, the focus should be on introducing digital tools and process innovations that help nurses work more efficiently. By implementing such approaches, healthcare systems can reduce nursing workload while preserving the high-quality care that patients already appreciate.

#### 4.1.2. Suggestions for future improvement

To address these issues, three potential solutions were identified from the interviews: (1) Self-reporting questionnaire, (2) Digital pre-admission system, (3) Enhanced EHR integration, and (4) User-centered system designs.

First, an important consideration is self-reporting questionnaire. While there is no established official questionnaire of nursing admission assessment in hospital, many general ward units already have been using their own questionnaire. Nurses reported having “too many tasks to complete,” which left them with insufficient time for thorough admission interviews. One key barrier with a self-reporting questionnaire is the interpretation of medical terminology. In the study hospital, many patients stated that some terms were too difficult to understand. While some patients asked for clarification of unfamiliar words to nurses or caregivers, others did not. This lack of understanding of medical terminology can result in misinformation being communicated to nurses, ultimately compromising patient safety. Therefore, when developing a self-reporting questionnaire, it is essential to address this issue with careful attention. The importance of using plain language is particularly emphasized in the development of patient-reported outcome measures (PROM) [41]. Adopting a patient-centered approach inspired by PROM methodologies in questionnaire design can add substantial value, ensuring clarity, accuracy, and improved patient engagement.

Second is the digital pre-admission system. We found that the appropriate timing for nursing admission data collection is three to seven days before patient admission. From the interviews, we established that bringing pre-admission data could relieve both nurses and caregivers by resolving uncertainty about patient information. While nurses were concerned about incoming patients’ conditions and what care they would need to provide, caregivers felt uncertain about which questions they would need to answer during admission. By utilizing a digital pre-admission system, these needs could be met. There were also demands for detailed information about what to prepare before admission from patients and caregivers. Providing this information along with the questionnaire would motivate and benefit all patients and caregivers. When developing the system, establishing a trustworthy and secure platform is crucial. This is particularly important as many elderly patients expressed concerns about voice phishing scams, which commonly target older adults.

Third, enhanced EHR integration is an essential feature. One of the burdens nurses reported during admission is documentation burden, which is a subsequent task after the questionnaire is completed. They must copy the answers into the EHR nursing admission assessment form, which creates additional work. However, this could be resolved by utilizing a mobile questionnaire that sync with the EHR system. Nurses unanimously agreed that the reliability of this mobile-collected data could be improved when nurses validate and sign off on it during its temporary storage.

Finally, user-centered system designs are paramount. Both usability and accessibility need to be carefully considered when designing digital solutions for nursing admission assessments. The interface should be intuitive enough for users of all technological proficiency levels while maintaining professional standards. Additionally, the system must incorporate features that accommodate various user needs, such as adjustable text sizes, voice input options, and clear navigation cues. To address security concerns, particularly among elderly patients who are wary of digital scams, the system should implement robust security measures and clearly communicate its official hospital affiliation, making the digital experience both safe and less daunting for stakeholders alike.

### 4.2. Broader Implications

Our investigation revealed that the roles and characteristics of caregivers for elderly patients largely reflect the broader social context, particularly in South Korea. As South Korea has become a super-aging society [42], the increasing need for elderly care has emerged [43]. However, South Korea is not the only country experiencing a super-aging society; other countries around the world are also undergoing demographic transitions [44]. In light of these global trends, the future of healthcare must prioritize integrated care models that address the needs of both patients and caregivers.

At the same time, healthcare systems worldwide are facing a chronic nursing shortage, even as the demand for care services continues to rise[45]. The COVID-19 pandemic has underscored the urgency of addressing this issue, reinforcing the importance of developing strategies to more effectively support nurses [46, 47]. A systematic review found multiple underlying factors contributing to nursing workforce shortages, including policy failures, education gaps, and high turnover rates[48]. One of the major contributor is the significant stress and burnout experienced by nurses, with excessive documentation burden cited as one of the key stressors.

Together, these challenges — the growing demands of an aging population, the increasing role of caregivers, and the global shortage of nurses — call for the development of innovative, integrated care strategies. Reducing the indirect nursing, such as documentation burden, and empowering both caregivers and nurses through supportive systems will be indispensable for sustaining healthcare delivery in aging societies.

### 4.3. Strengths and Limitations

A major strength of this study is its integration of diverse stakeholder perspectives—nurses, patients, and caregivers—on the nursing admission assessment process. By conducting interviews across different participant groups, the study captured a comprehensive understanding of the real-world challenges and needs in clinical settings. This methodology provides valuable insights for developing patient-centered and caregiver-integrated approaches to nursing workflows. Furthermore, our findings contribute to the broader discourse on digital transformation in healthcare, highlighting the importance of demand-driven innovation rather than supply-driven adoption. In particular, intensive interviews with nurses provided in-depth insights into the entire nursing process, which is crucial for the development of healthcare application[49]. By understanding the complex dynamics among healthcare endusers and their interactions, hospitals can design more inclusive and effective digital solutions that maintain care quality while addressing emerging workforce challenges. However, this study has several limitations. It was conducted at a single tertiary hospital in South Korea, which may limit the generalizability of findings to different institutional contexts or demographic groups of nurses, patients, and caregivers. The hospital’s strong brand reputation could have introduced positive response bias, as many patients expressed great satisfaction during interviews, potentially masking dissatisfaction with the admission process. Although the researchers were not involved in direct patient care and emphasized confidentiality, the fact that they were registered nurses affiliated with the study hospital might have made patients hesitant to voice criticisms openly. While efforts were made to mitigate these biases, their potential influence on the findings cannot be entirely excluded. Despite these limitations, the study offers valuable insights into the real-world challenges and opportunities for improving nursing admission assessments, particularly within large tertiary care contexts. Future studies should aim to validate these findings across diverse healthcare settings.

## 5. Conclusion

This study uncovered critical unmet needs and challenges within the nursing admission assessment process by integrating the perspectives of nurses, patients, and caregivers. Time pressures, documentation burdens, and communication barriers were found to significantly affect both the quality and efficiency of nursing assessments. To address these issues, we recommend the development of a digital preadmission system designed with a strong focus on user-centered features, aimed at streamlining workflows and enhancing care quality. Future research should prioritize the implementation and evaluation of such systems to ensure they effectively reduce nurses’ workloads while maintaining the high standards of patient care and safety that are essential in clinical practice.

## Supporting information

COREQ Checklist

Interview guide for RN

Interview guide for patients and caregivers

Coding Framework

## Data Availability

The data supporting the findings of this study are contained in the manuscript; raw interview data are not publicly available to protect participant confidentiality.

## CRediT authorship contribution statement

Choi, Y.Y.: Writing – review & editing, Investigation, Methodology, Conceptualization, Project administration, Formal analysis.

Seo, J.H.: Writing – original draft, Investigation, Methodology, Conceptualization, Project administration, Formal analysis.

Shin, H.A.: Writing – review & editing, Investigation, Formal analysis.

Park, M.J: Writing – review & editing, Formal analysis.

Lee, Y.E: Writing – review & editing, Formal analysis.

Jeon, S.B: Writing – review & editing, Formal analysis.

Lee, M.J.: Writing – review & editing, Investigation, Project administration.

Park, S.H.: Writing – review & editing, Project administration, Supervision, Methodology, Funding acquisition, Conceptualization.

Hong, J.H.: Writing – review & editing, Supervision, Methodology, Funding acquisition, Conceptualization.

## Funding

This study was supported by the research fund of the Department of Nursing, Samsung Medical Center in 2025 (No. SMC-NSD-2025-01)

## Declaration of competing interests

The authors declare no competing interests.

## Acknowledgments

We express our sincere gratitude to nursing managers of both 08E and C6W wards at Samsung Medical Center for their cooperation in our study.

## Notes

### Competing Interest Statement

The authors have declared no competing interest.

### Author Declarations

IRB of Samsung Medical Center gave ethical approval for this work

