## Supplementary material for "Understanding Unmet Needs and Opportunities for Digital Transformation in Nursing Admission Assessment: A Qualitative Study": COREQ Checklist

### Supplementary File 1

#### COREQ Checklist

| COREQ Checklist | Reported in Manuscript |
|---|---|
| **Domain 1: Research team and reflexivity** | |
| *Personal Characteristics* | |
| 1. Interviewer/facilitator | Yes. Subsection 2.2 Data Collection |
| 2. Credentials | Yes. CRediT authorship contribution statement |
| 3. Occupation | Yes. Registered Nurses |
| 4. Gender | Yes. Females (YC, HS) and one male (JS) |
| 5. Experience and training | Yes. Subsection 2.2 Data Collection |
| *Relationship with participants* | |
| 6. Relationship established | Yes. Some participants may have known members of the research team from the same hospital |
| 7. Participant knowledge of the interviewer | Yes. Some participants knew researchers' names and roles |
| 8. Interviewer characteristics | Yes. Stated on the Title Page |
| **Domain 2: Study design** | |
| *Theoretical framework* | |
| 9. Methodological orientation and theory | Yes. Subsection 2.1 Study Design |
| *Participant selection* | |
| 10. Sampling | Yes. Subsection 2.3 Participants and Recruitment |
| 11. Method of approach | Yes. Subsection 2.3 Participants and Recruitment |
| 12. Sample size | Yes. Subsection 2.3 Participants and Recruitment |
| 13. Non-participation | No. No one refused |
| *Setting* | |
| 14. Setting of data collection | Yes. Subsection 2.2 Data Collection |
| 15. Presence of non-participants | No. Only participants and researchers were present |
| 16. Description of sample | Yes. Subsection 3.1 Participant Characteristics |
| *Data collection* | |
| 17. Interview guide | Yes. Supplementary File 2, 3 |
| 18. Repeat interviews | No |
| 19. Audio/visual recording | Yes. Subsection 2.2 Data Collection |
| 20. Field notes | Yes. Subsection 2.2 Data Collection |
| 21. Duration | Yes. Subsection 2.2 Data Collection |
| 22. Data saturation | Yes. Result section |
| 23. Transcripts returned | Yes for nurses; not for patients & caregivers (discharged) |
| **Domain 3: Analysis and findings** | |
| *Data analysis* | |
| 24. Number of data coders | Yes. Subsection 2.3 Data Analysis |
| 25. Description of the coding tree | Not reported |

| **COREQ Checklist** | **Reported in Manuscript** |
|---|---|
| 26. Derivation of themes | Not reported |
| 27. Software | Yes. MAXQDA (v24.8.0) |
| 28. Participant checking | Yes for nurses; not for patients & caregivers (discharged) |
| *Reporting* | |
| 29. Quotations presented | Yes. Result section |
| 30. Data and findings consistent | Yes. Findings consistent with data |
| 31. Clarity of major themes | Yes. Clearly described |
| 32. Clarity of minor themes | Yes. Subsection 3.7 Concerns |
