## Supplementary material for "Understanding Unmet Needs and Opportunities for Digital Transformation in Nursing Admission Assessment: A Qualitative Study": Interview guide for RN

### Supplementary File 2

#### Interview Guide for Registered Nurses

This interview guide was used for focus group interviews (FGIs) with registered nurses to explore their experiences with the nursing admission assessment process.

#### 1. Experience with the Current Nursing Admission Assessment Process

**Main Question:**

Could you describe how the current nursing admission assessment process is conducted, including preparation, execution, and documentation?

##### (1) Time Required

- On average, how long does it take to prepare for, conduct, and document the nursing admission assessment? What factors influence whether it takes longer or shorter?
- When during the admission process do you usually perform the assessment? What are the reasons or criteria for this timing?
- How does the assessment process impact the time you spend on direct patient care? If direct care time is reduced, how does this affect your overall work?

##### (2) Workload Burden

- How significant is the nursing admission assessment within your overall workload? Have you ever felt burdened by it? If so, in what situations?
- What aspects of the assessment process are most challenging? What support or changes would help alleviate these difficulties?
- When patient information is lacking at admission, what challenges do you face? How could the process be improved to manage these situations more efficiently?

##### (3) Patient Perspective on the Assessment

- With whom do you usually conduct the assessment – the patient, caregiver, or both? Are there differences in how you interact depending on the respondent?
- Do you feel that patients are given enough opportunity to share their stories? If not, what challenges have you encountered?
- What factors make it difficult for patients to fully share their information? What improvements could help address this?
- What changes to the nursing admission assessment process would help gather more accurate and complete information from patients? (e.g., question style, environment, time allocation)

#### 2. Nursing Admission Assessment Items and Additional Questions

**Main Question:**

Do you think the current nursing admission assessment items adequately reflect the patient's health status? If not, in what areas do you feel improvements are needed?

##### (1) Appropriateness of Current Items

- Are there any items you find unnecessary or redundant? If so, which ones, and how could they be improved or replaced?
- Are there any aspects of a patient's health that are not currently addressed but should be included? Why do you think these additions would be helpful?
- Do you feel the current assessment items adequately reflect the characteristics of different units (e.g., medical, surgical wards)? If not, what additional items or modifications would you suggest?
- Are there items or expressions that patients often find difficult to understand? What improvements would make these easier for patients to comprehend?

**(2) Challenges Related to Additional Questions**

- Have you ever felt the need to ask additional questions beyond the standard items during the assessment? (e.g., asking about MRI compatibility for patients with implanted devices) If so, what kinds of questions were needed, and what challenges did you encounter?
- After completing an assessment, have you ever realized that an important question was omitted? If so, what types of questions were missing, and how could the process be improved to avoid such omissions?
- What support or tools do you think nurses need to effectively ask additional questions? (e.g., structured forms, digital tools)
- Do you think the use of additional questions varies depending on the nurse's experience level? If so, why, and what systems or tools could help standardize the process?

#### 3. Recommendations for future improvement

Do you have any additional suggestions regarding the improvement of the nursing admission assessment?
