## Supplementary material for "Understanding Unmet Needs and Opportunities for Digital Transformation in Nursing Admission Assessment: A Qualitative Study": Interview guide for patients and caregivers

### Supplementary File 3

#### Interview Guide for Patients and Caregivers

This interview guide was used for in-depth interviews with patients and caregivers to explore their experiences with the nursing admission assessment process.

#### 1. Experience with Nursing Information Collection

**Main Question:**

When you were admitted, do you recall the experience of a nurse asking about your health status? Please share any thoughts or feelings you had during that process.

##### (1) Time and Communication

- How did you feel about the speed at which the nurse asked questions? (e.g., Did it feel too fast, appropriate, or too slow? If it felt inappropriate, how would you have preferred it?)
- Were the questions easy to understand? Were there any topics you wished the nurse had explored further?
- Did you feel that the nurse listened attentively to your story? If not, what made you feel that way?
- Were there moments when you wanted to say more but couldn't? If so, in what situations did this occur?

##### (2) Privacy

- Were you concerned that personal information might be overheard by caregivers or other patients nearby? If so, can you describe the situation?
- Were there any questions that felt uncomfortable or difficult to answer? What were they, and why did you find them difficult?

#### 2. Experience with Self-Reported Method

**Main Question:**

How did you feel about recording your health status by filling out a paper form yourself? Please share any convenient or inconvenient aspects you experienced while completing it.

##### (1) Difficulties with Self-Reporting

- How did the experience differ between answering questions asked by a nurse and filling out the form yourself?
- What aspects were the most challenging when completing the paper form? (e.g., difficulty writing, difficulty understanding the questions)
- Were there any questions or expressions on the paper form that were difficult to understand? Please describe any examples you recall.
- When you encountered difficult questions while completing the form, how did you usually handle them? (e.g., asking a nurse for help, skipping the question, guessing the answer)

#### 3. Opinions on Future Self-Reporting Methods

**Main Question:**

What do you think about using electronic devices such as mobile phones or tablets to record your health status instead of using paper forms? Do you think this method would be convenient or difficult?

##### (1) Use of Electronic Devices

- What aspects might be inconvenient when using a mobile phone or tablet to complete a questionnaire? (e.g., small text size, difficulty entering information)
- Among input methods (pressing buttons, typing text, using voice input), which would you find most convenient?
- If there were features to assist when questions are difficult to answer, what would be most helpful? (e.g., simple explanations, examples, images, voice guidance)

#### 4. Opinions on the Timing of Self-Reporting

**Main Question:**

If you could complete the self-report questionnaire before admission, how do you think it would help you?

##### (1) Timing and Information Needs

- If you were to complete the questionnaire before admission, how many days in advance would be appropriate? Please explain why you think that timing would be suitable.
- If there was information you would have liked to receive before admission, what would have been helpful? (e.g., items to prepare for hospitalization, admission procedures)

#### 5. Additional Comments

Is there anything else you would like to share regarding your experience with the nursing admission assessment?
