## Supplementary material for "Understanding Unmet Needs and Opportunities for Digital Transformation in Nursing Admission Assessment: A Qualitative Study": Coding Framework

### Supplementary File 4

#### Coding Framework

| Theme | Sub-themes | Codes |
|---|---|---|
| Perceived meaning of Nursing Admission Assessment | Data-driven and comprehensive assessment | • Pre-admission chart review<br>• Integration of outpatient records<br>• Clinical reasoning based on data interpretation |
| | Starting point of holistic care | • Comprehensive head-to-toe assessment<br>• Continuity from admission to discharge<br>• Recognition of assessment as essential |
| Operational and systemic challenges in Nursing Admission Assessment | Managing time pressure with a self-report questionnaire | • Time pressure in assessments<br>• Reliance on patient a self-report questionnaire<br>• Workflow disruptions from incomplete questionnaires<br>• Pre-filling assessments based on EHR records<br>• Struggles of patients and caregivers to provide timely information |
| | Emotional burden under unpredictable admissions | • Unpredictable admissions<br>• Rushed assessments<br>• Nurse guilt and frustration<br>• Patient-perceived satisfaction |

| Theme | Sub-themes | Codes |
|---|---|---|
| | Documentation burden and systemic inefficiencies | • Loss of direct nursing care time<br>• Burden of repetitive documentation for each admission<br>• Lack of interoperability across hospitals<br>• Time-consuming medication reconciliation |
| Communication challenges among stakeholders | Challenges in communicating with elderly patients | • Communication barriers with elderly patients<br>• Memory issues in recalling medical history<br>• Hearing and vision impairments |
| | Essential role of caregivers | • Caregivers as crucial information sources<br>• Caregiver-dependent information accuracy<br>• Caregiver-assisted efficiency |
| | Misunderstandings of medical terminology and questionnaire items | • Misunderstanding of medical terms<br>• Need for translation into patient-friendly language<br>• Random responses to unclear items<br>• Increased assessment burden |
| Suggestions for future improvement | Digital pre-admission system | • Advance input of health information<br>• Linkage to EHR to facilitate assessment<br>• Positive patient and caregiver perception<br>• Mobile-based questionnaire adoption |

| Theme | Sub-themes | Codes |
|---|---|---|
| | System design accommodating stakeholders' needs | • Separation of patient-input and nurse-assessed items<br>• Need for nursing verification of patient-entered data<br>• Use of patient-friendly terminology<br>• Enhancing visual accessibility<br>• Offering flexible input options |
| | Concerns about digital transformation of Nursing Admission Assessment | • Risk of over-detailed patient input<br>• Resistance to new digital systems<br>• Patient anxiety about errors and technology<br>• Preference for direct human interaction |
